# Large Language Model Symptom Identification from Clinical Text: A Multi-Center Study

**DOI:** 10.1101/2024.12.16.24319044

**Authors:** Andrew J. McMurry, Dylan Phelan, Brian E. Dixon, Alon Geva, Daniel Gottlieb, James R. Jones, Michael Terry, David Taylor, Hannah Grace Callaway, Sneha Mahoharan, Timothy Miller, Kenneth D. Mandl

**Author notes:** Corresponding Author: Kenneth D. Mandl Computational Health Informatics Program, Boston Children’s Hospital, 401 Park Drive, LM5506, Mail Stop BCH3187, Boston, MA 02215.

## Abstract

Recognition of patient symptoms is core to medicine, research, and public health. We tested four large language models (LLMs) identifying 11 symptoms of infectious respiratory diseases from emergency department notes (N=204). Each LLM outperformed ICD-10-based identification. GPT-4 had highest tested accuracy, F1 score 91.4% vs. 45.1% for ICD-10. GPT-4 performance in an independent validation cohort (N=308) was even higher with an F1 score of 94.0%.

## Main text

To practice medicine, accurate identification and interpretation of symptoms are paramount. Symptoms are primary indicators of a patient’s health state, underpinning diagnostic processes^1^ and choice of therapeutic interventions.^2^ Identifying symptoms is fundamental for matching patients to clinical trials, and conducting public health surveillance.^3–6^ While routinely documented in physician notes, coded formats often underreport present symptoms.^4,7–10^ We sought to measure accuracy of state-of-the-art commercial and open source large language models (LLMs) for symptom identification, with a focus on infectious respiratory disease symptoms.^4^

Legacy natural language processing (NLP) systems have struggled with key aspects of symptom interpretation, including recognizing uncertainty,^11^ determining relevance of a symptom to the current encounter,^12^ and discerning temporality.^13^ The context^10,14,15^ surrounding infectious respiratory diseases includes symptoms pertaining to acute infections, noninfectious conditions, treatment side effects,^6^ indications for treatment, or patient instructions (e.g., “Use albuterol inhaler as needed for difficulty breathing.”). Newer LLMs may not require localized training or domain-specific adaptation^16–18^ to achieve high performance in this domain.

We evaluated the performance of four LLMs (GPT-4 turbo,^19^ GPT-3.5-turbo,^19^ Llama2-70B,^20^ and Mixtral 8×7B^21^) when tasked to emulate human chart reviewers and detect symptoms in emergency department (ED) notes from the electronic health record (EHR). Manual chart review was performed to annotate ground truth labels. LLMs were provided “zero-shot” chart review prompt instructions without AI model training or fine tuning. Eleven infectious disease symptoms related to acute respiratory disease were evaluated: congestion or runny nose, cough, diarrhea, dyspnea (shortness of breath), fatigue, fever or chills, headache, loss of taste or smell, muscle or body aches, nausea or vomiting, and sore throat.

Accuracy for each LLM was considerably higher for all infectious disease symptom identification tasks compared to conventional ICD-10 based identification. Results for all four LLMs are presented in Supplementary tables. In our Test cohort at Site 1—a large northeastern urban pediatric academic medical center—GPT-4 accuracy was the highest, F1=91.8% compared to the ICD-10 based detection F1=45.1%. Accuracy of ICD-10 was a comparator based on published data;^4^ lower performance scores for symptom identification is consistent with prior literature.^8,10^ GPT-4 was then validated in an independent cohort at Site 2—a midwestern state-wide health information exchange—without any further model training or fine tuning. Performance was improved in this validation cohort with F1=94.0%. Impressively, GPT-4 was more transferable from Site 1 to Site 2 (2.6% absolute increase F1-score) than ICD-10 (18.2% absolute decrease F1 score). Notably, ICD-10 coding practices across healthcare settings vary considerably.^22^ GPT-4 accuracy was higher than ICD-10 based methods for all symptoms across both sites (Figure 1).

**Figure 1.**
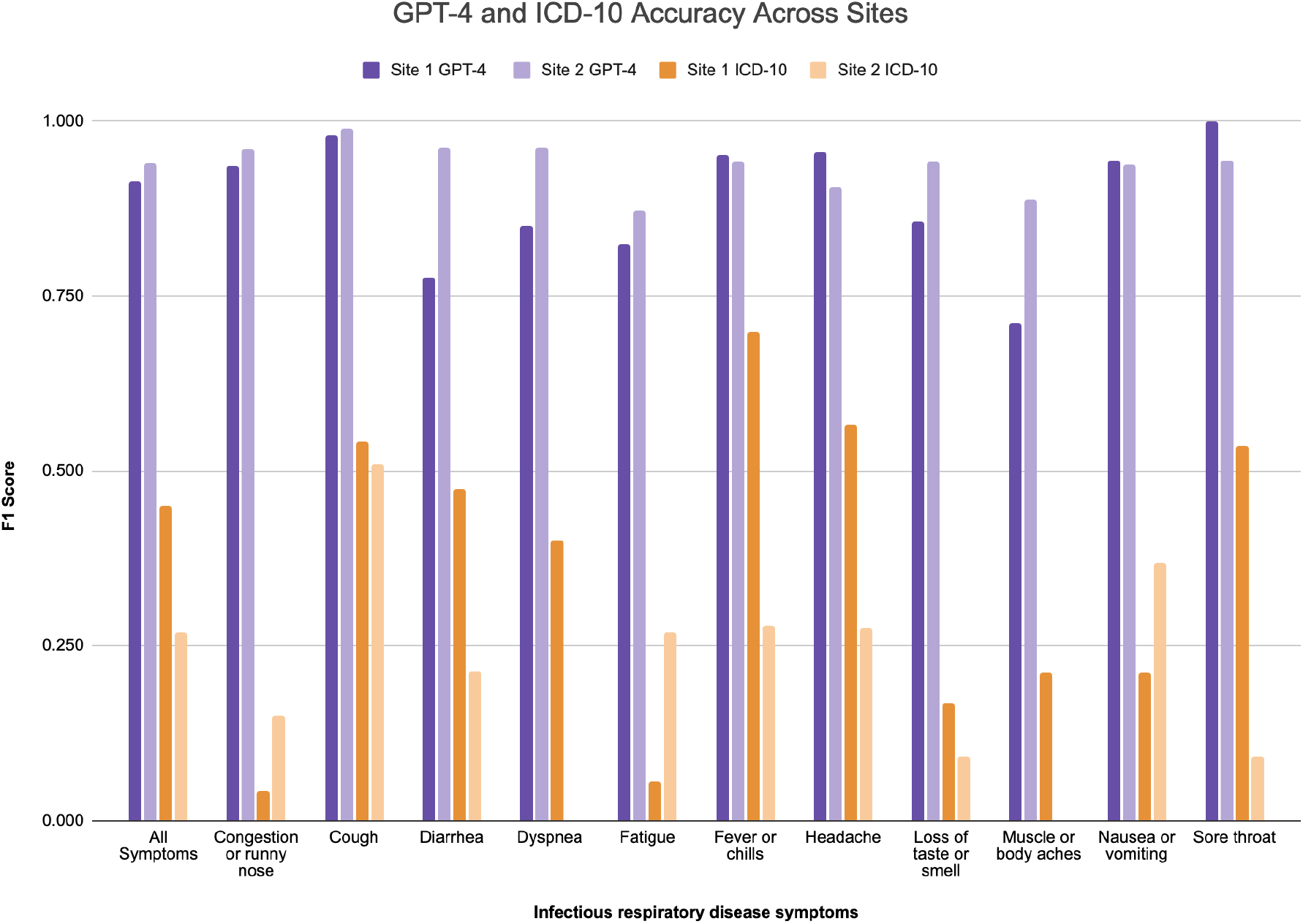
Symptom detection accuracy of GPT-4 versus ICD-10 at two healthcare sites. Site 1 is a large northeastern urban pediatric academic medical center. Site 2 is a midwestern state-wide health information exchange. F1 scores are shown for each method and symptom benchmarked against ground truth labels from chart reviews in Test and Validation cohorts.

This study highlights the transformative potential of large language models (LLMs) in accurately identifying symptoms from clinical text, addressing long standing challenges in medicine and public health. However, important limitations merit consideration. The evaluation focused specifically on symptoms related to infectious respiratory diseases. While this represents a critical use case, the generalizability of LLMs to other clinical domains and broader symptom categories (i.e., too numerous to include in an LLM prompt) remains to be validated. Additionally, while GPT-4 demonstrated strong transferability between two healthcare settings, including a statewide HIE encompassing 38 distinct health systems with highly heterogeneous data and coding practices, its zero-shot approach could face challenges in environments with institution-specific documentation practices. However, the diversity of our setting suggests it is well-suited to handle the variability inherent in human expression of language and concepts, which may pose challenges for more rigid approaches such as those relying solely on ICD coding.

Scaling to larger, more diverse cohorts—beyond ED populations and incorporating automated benchmarking techniques—could strengthen the reliability of assessments. Furthermore, although ICD-10 coding was used as a comparator, it is a limited benchmark given its well-documented shortcomings in capturing clinical nuance.^22^ Traditional NLP tools like Apache cTAKES^23^ or Metamap^24^ can extract mentions of symptoms in notes by matching against terminologies but cannot be directly compared because they require post-processing steps to produce note-level extractions. Our findings underscore the potential of LLMs to address gaps in traditional symptom identification methods, paving the way for advancements in diagnosis, clinical trial matching, and surveillance. Future research should focus on refining these models to ensure equitable, efficient, and seamless integration into diverse healthcare settings.

## Methods

### Study Design

This is a retrospective study to identify acute respiratory infection symptoms from clinical text. In a Development cohort, at a single site, prompting strategies were developed for open source models Llama 2 70B Chat and Mistral AI’s Mixtral 8×7B Instruct model as well as commercial models from GPT-3.5 turbo (version 0125) and GPT-4 turbo (version 0125). After identifying the best performing prompting strategies for each LLM, performance across models was evaluated using a Test cohort at the same site and compared against ICD-10 based methods. The top performing model was then executed on an independent Validation cohort in a different setting.

### Setting and cohorts

Site 1 is a large northeastern urban pediatric academic medical center. Site 2 is a midwestern statewide health information exchange network that includes 120 EDs. The cohorts comprised patients who visited the ED between March 1, 2020 and May 31, 2022. At Site 1, patients 21 years old and younger were included. At Site 2, patients of any age who had a COVID-19 diagnosis at any time during the study period were included. Cohorts were constructed through sampling to ensure that 30 mentions of each symptom were present. The Development cohort (N=103) from Site 1 was created for prompt engineering. A Test cohort (N=204) from Site 1 was for model evaluation. The Validation cohort (N=308) from Site 2 was for evaluating transferability of the model. Analyses were conducted in a secure environment including a HIPAA-compliant Azure GPT-4 instance.

The Boston Children’s Hospital Committee on Clinical Investigation (BCH IRB-P00043392) and the Indiana University institutional review board (IU IRB 24673) both determined the study to be exempt from full human subjects oversight. Waivers of consent were obtained to allow the targeted extraction and secure review of clinical notes by approved study personnel within protected environments.

### Measures

Metrics include micro F1 score, precision and recall, evaluated against ground truth labels across 11 symptoms. All metrics were calculated for each symptom and for all symptoms together.

### Prompt engineering

For each model, prompting strategies^25^ were developed to mirror human clinical annotation guidelines. Prompts ranged in complexity from an identity prompt, wherein LLMs were instructed to assume the identity of a chart reviewer, to a verbose prompt containing symptom specific criteria. Figure 2 summarizes the five types of prompt instructions attempted. LLM generated responses were then combined with four output parsing pipelines for a total of twenty prompting strategies. LLM generated responses were parsed into structured data containing boolean results denoting symptom presence or absence. Structured responses are compared against ground truth labels to measure accuracy for each LLM in each prompt strategy. Results are compared to identify each LLM’s optimal prompting strategy (Supplementary tables).

**Figure 2:**
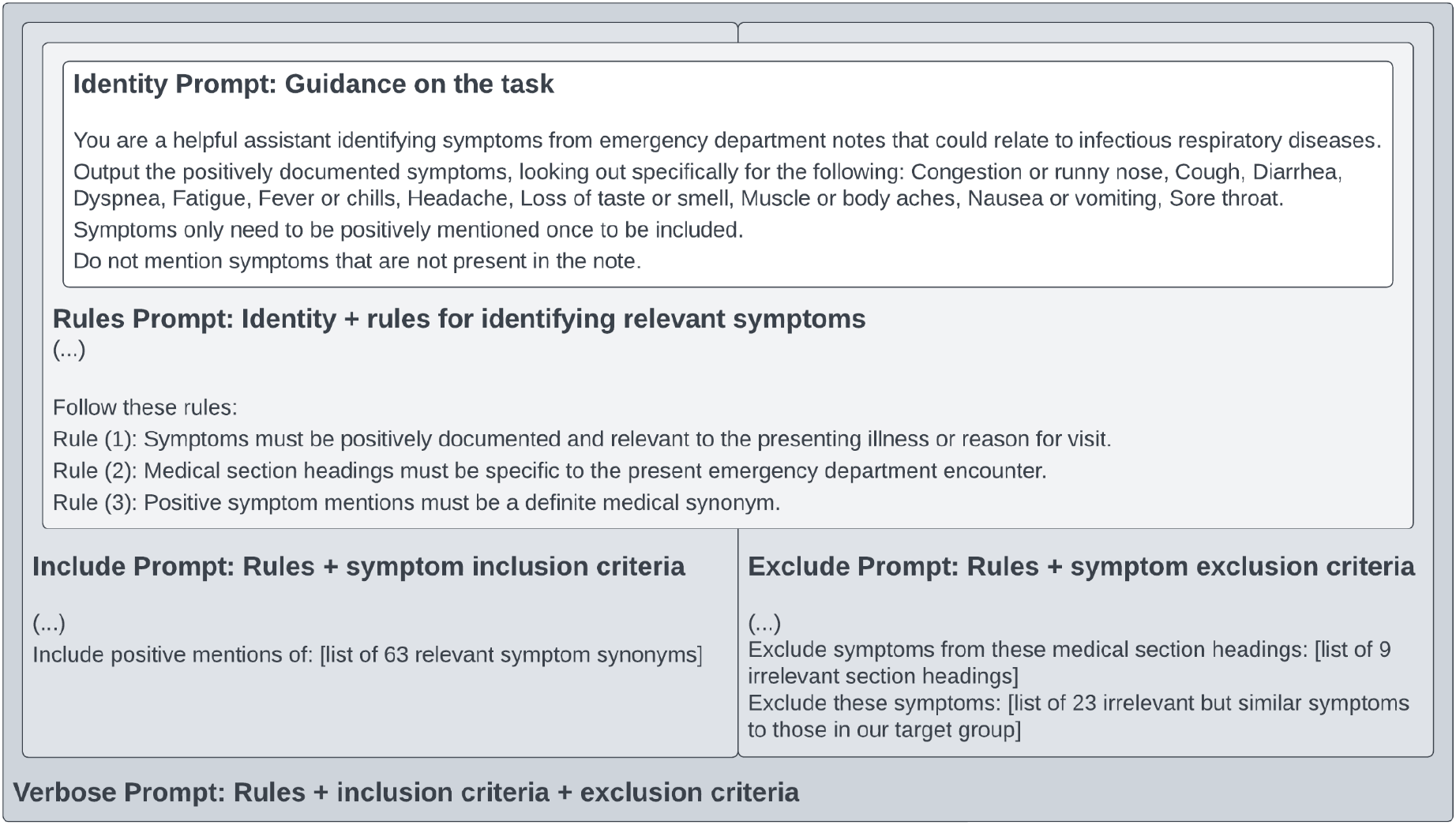
LLM Prompts intended to reproduce chart review criteria. The identity prompt contains text present in every type of prompt. The rules prompt extends the identity prompt with basic chart review guidelines. Include and exclude prompts extend the rules prompt with symptom specific criteria. The verbose prompt combines all prompts to approximate the same chart review criteria as human subject matter experts.

### Comparator

ICD-10 code sets for each symptom were previously defined by subject matter experts.^4^ LLM performance was compared with previously published ICD-10-based symptom identification performance^4^ at Site 1 and against new ICD-10-based measurements at Site 2. Ground truth labels were compared to ICD-10 and LLM based methods for symptom identification. ICD-10 codes included any ICD-10 code recorded during the ED encounter at the time of ED discharge.^4^

## Data availability

Supplementary tables and supplementary figures contain results of LLM prompts and ICD10 methods for each cohort evaluated.

## Code availability

All code including LLM prompts and results are freely available at https://github.com/smart-on-fhir/infectious-symptoms-llm-study.

## Acknowledgements

Support for this study was provided by the Advanced Research Projects Agency for Health (ARPA-H) (75N95023D00001, 75N95023F00019), the National Center for Advancing Translational Sciences (NCATS), National Institutes of Health (U01TR002623), the Office of the National Coordinator for Health Information Technology (ONC) (90AX0031, 90C30007), and the the Centers for Disease Control and Prevention (CDC) of the US Department of Health and Human Services as part of a financial assistance award.

## Author contributions

Conceptualization (KDM, AJM, DP, AG, TM, JJ, DG), Data Curation (AJM, AG, HGC, SM), Formal Analysis (AJM, DP ), Funding Acquisition (KDM), Investigation (AJM, DP, AG, DT, HGC, SM), Methodology (KDM, AJM, DP, DG, TM, JJ), Project Administration (JJ, BD, DT), Software (DP, AJM, MT, DG), Supervision (KDM, TM), Validation (BD, DT, HGC, SM), Visualization (DP, JJ), Manuscript Preparation (AJM, DP, KDM), Review and Editing (All authors).

## References

1. Chen, A., Chen, D. O. & Tian, L. Benchmarking the symptom-checking capabilities of ChatGPT for a broad range of diseases. J. Am. Med. Inform. Assoc. (2023) doi:10.1093/jamia/ocad245.

2. Harskamp, R. E. & De Clercq, L. Performance of ChatGPT as an AI-assisted decision support tool in medicine: a proof-of-concept study for interpreting symptoms and management of common cardiac conditions (AMSTELHEART-2). Acta Cardiol. 79, 358–366 (2024).

3. Mandl, K. D. et al. Push Button Population Health: The SMART/HL7 FHIR Bulk Data Access Application Programming Interface. NPJ Digit Med 3, 151 (2020).

4. McMurry, A. J. et al. Moving Biosurveillance Beyond Coded Data Using AI for Symptom Detection From Physician Notes: Retrospective Cohort Study. J. Med. Internet Res. 26, e53367 (2024).

5. Matheny, M. E. et al. Enhancing Postmarketing Surveillance of Medical Products With Large Language Models. JAMA Netw Open 7, e2428276 (2024).

6. Henry, S., Buchan, K., Filannino, M., Stubbs, A. & Uzuner, O. 2018 n2c2 shared task on adverse drug events and medication extraction in electronic health records. J. Am. Med. Inform. Assoc. 27, 3–12 (2020).

7. Malden, D. E. et al. Natural Language Processing for Improved Characterization of COVID-19 Symptoms: Observational Study of 350,000 Patients in a Large Integrated Health Care System. JMIR Public Health Surveill 8, e41529 (2022).

8. Crabb, B. T. et al. Comparison of international classification of Diseases and Related Health Problems, Tenth Revision codes with electronic medical records among patients with symptoms of Coronavirus disease 2019. JAMA Netw. Open 3, e2017703 (2020).

9. Koleck, T. A., Dreisbach, C., Bourne, P. E. & Bakken, S. Natural language processing of symptoms documented in free-text narratives of electronic health records: a systematic review. J. Am. Med. Inform. Assoc. 26, 364–379 (2019).

10. Hardjojo, A. et al. Validation of a Natural Language Processing Algorithm for Detecting Infectious Disease Symptoms in Primary Care Electronic Medical Records in Singapore. JMIR Med Inform 6, e36 (2018).

11. Harkema, H., Dowling, J. N., Thornblade, T. & Chapman, W. W. ConText: an algorithm for determining negation, experiencer, and temporal status from clinical reports. J. Biomed. Inform. 42, 839–851 (2009).

12. Pugliese, G. et al. Are artificial intelligence large language models a reliable tool for difficult differential diagnosis? An a posteriori analysis of a peculiar case of necrotizing otitis externa. Clin Case Rep 11, e7933 (2023).

13. Miller, T., Bethard, S., Dligach, D. & Savova, G. End-to-end clinical temporal information extraction with multi-head attention. Proc Conf Assoc Comput Linguist Meet 2023, 313–319 (2023).

14. Wang, H., Gao, C., Dantona, C., Hull, B. & Sun, J. DRG-LLaMA : tuning LLaMA model to predict diagnosis-related group for hospitalized patients. NPJ Digit Med 7, 16 (2024).

15. Zhang, F., Laish, I., Benjamini, A. & Feder, A. Section Classification in Clinical Notes with Multi-task Transformers. in Proceedings of the 13th International Workshop on Health Text Mining and Information Analysis (LOUHI) (eds. Lavelli, A. et al.) 54–59 (Association for Computational Linguistics, Abu Dhabi, United Arab Emirates (Hybrid), 2022).

16. Lin, C. et al. Does BERT need domain adaptation for clinical negation detection? J. Am. Med. Inform. Assoc. 27, 584–591 (2020).

17. Zhou, W., Dligach, D., Afshar, M., Gao, Y. & Miller, T. A. Improving the Transferability of Clinical Note Section Classification Models with BERT and Large Language Model Ensembles. Proc Conf Assoc Comput Linguist Meet 2023, 125–130 (2023).

18. Nori, H. et al. Can Generalist Foundation Models Outcompete Special-Purpose Tuning? Case Study in Medicine. arXiv [cs.CL] (2023).

19. GPT-4. https://openai.com/index/gpt-4-research/.

20. Meta Llama 2. Meta Llama https://llama.meta.com/llama2/.

21. Mistral, A. I. Mixtral of experts. https://mistral.ai/news/mixtral-of-experts/ (2023).

22. Nelson, S. J. et al. Are ICD codes reliable for observational studies? Assessing coding consistency for data quality. Digit. Health 10, 20552076241297056 (2024).

23. Savova, G. K. et al. Mayo clinical Text Analysis and Knowledge Extraction System (cTAKES): architecture, component evaluation and applications. J. Am. Med. Inform. Assoc. 17, 507–513 (2010).

24. Aronson, A. R. & Lang, F.-M. An overview of MetaMap: historical perspective and recent advances. J. Am. Med. Inform. Assoc. 17, 229–236 (2010).

25. Wang, L. et al. Prompt engineering in consistency and reliability with the evidence-based guideline for LLMs. NPJ Digit Med 7, 41 (2024).

