## Supplementary figures for "Large Language Model Symptom Identification from Clinical Text: A Multi-Center Study"

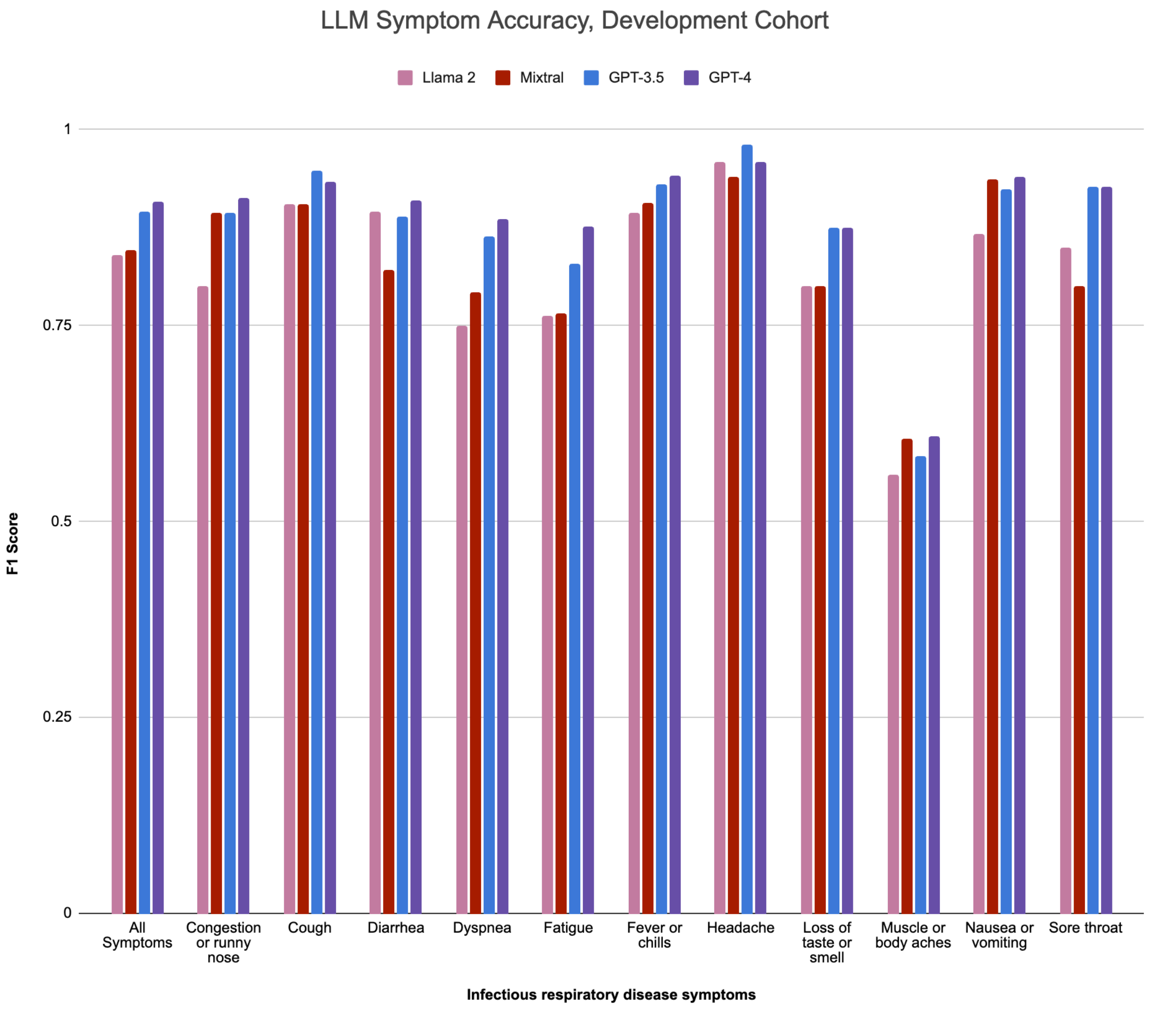


**Figure S1. F1 scores for the best strategy using the development cohort at Site 1.** Each color denotes a symptom identification method: Llama2, Mixtral, GPT-3.5, and GPT-4. Each of 11 infectious disease symptoms are shown as well as summary score for all symptoms. Overall, GPT-4 performed best with micro F1-score 90.8% for all symptoms.

Large Language Model Symptom Identification from Clinical Text: A Multi-Center Study

McMurry AJ, Phelan D, Dixon BE, Geva A, Gottlieb D, Jones JR, Terry M, Taylor D, Callaway HG, Mahoharan S, Miller T, Mandl KD.

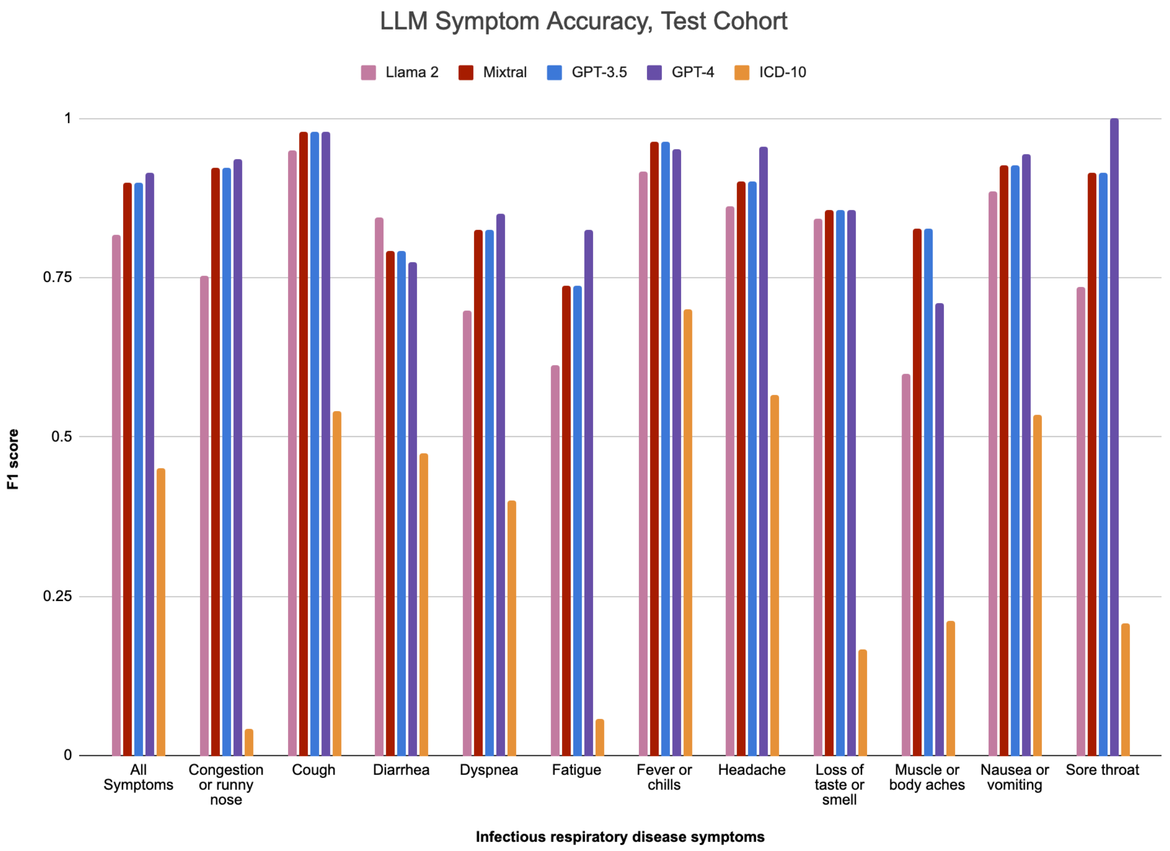


**Figure S2: F1 scores for the best strategy using the test cohort at site 1.** Each color denotes a symptom identification method: Llama2, Mixtral, GPT-3.5, GPT-4, and ICD-10. Each of 11 infectious disease symptoms are shown as well as  score for all symptoms. Overall, GPT-4 performed best with micro F1-score 91.4% for all symptoms.
